# Real-World External Validation of a Clinically Implemented Artificial Intelligence Index for Visual Field Severity from Optical Coherence Tomography

**DOI:** 10.64898/2026.09.08.26362248

**Authors:** Yukiko Kora, Hiromi Kora, Toshitake Kora

## Abstract

**Purpose:** To externally evaluate the unitless optical coherence tomography–derived AI-CO index for association with Humphrey mean deviation (MD) and discrimination of MD-defined visual field severity.

**Methods:** This retrospective study included 819 eyes of 424 patients receiving glaucoma care, with AI-CO analysis and Humphrey 24-2 SITA Faster or 30-2 SITA Fast testing within 365 days. The unitless output labeled “peripheral” was termed AI-CO all. Primary analyses assessed Spearman rank correlation and discrimination at MD thresholds of −2, −6, and −12 dB. Additional analyses assessed adjusted associations and one-eye-per-patient and program-specific robustness. Exploratory analyses assessed nonlinearity and cutoffs with patient-level cross-validation.

**Results:** AI-CO all correlated with Humphrey MD (Spearman ρ=0.785; patient-cluster 95% confidence interval [CI], 0.750–0.818). Median AI-CO all values decreased from −2.10 in the normal range to −4.40, −10.15, and −17.35 in mild, moderate, and severe categories, respectively (ordered trend P<0.001). Areas under the receiver operating characteristic curve were 0.870 (95% CI, 0.842–0.895), 0.919 (0.894–0.939), and 0.943 (0.920–0.963) at the three thresholds. The adjusted spline detected modest nonlinearity with little gain in explained variance over a linear model (R^2^, 0.735 to 0.738). Associations remained after covariate adjustment, and findings were similar in the one-eye-per-patient analysis and across visual field programs.

**Conclusions:** AI-CO all showed a rank correlation with Humphrey MD and differentiated eyes across specified MD thresholds. It should be interpreted as a severity-related index that complements, rather than replaces, measured perimetry. Exploratory AI-CO all cutoffs require independent validation.

**Translational Relevance:** In routine glaucoma care, AI-CO all showed a rank correlation with Humphrey MD and discriminated MD-defined severity.

## Introduction

Standard automated perimetry remains essential for evaluating visual function in glaucoma and other optic nerve disorders. Its results, however, depend on patient understanding and sustained responses and may be influenced by test duration, fatigue, learning effects, fixation stability, and response reliability. Optical coherence tomography (OCT) provides rapid and objective assessment of retinal structure, but translating structural findings into clinically interpretable functional severity remains challenging.

Deep-learning models have estimated pointwise visual field sensitivity and global visual field indices from retinal thickness maps or unsegmented OCT images^1,2^. Other models have estimated the central 10-degree visual field from OCT images, sometimes incorporating 24-2 or 30-2 information^3–5^. Koyama et al. more recently described a segmentation-free three-dimensional convolutional neural network that estimated both 24-2 and 10-2 visual fields directly from OCT images^6^. These studies established technical feasibility, but most evaluated development or research datasets. They provide limited evidence regarding a commercially implemented system used in routine care, where image acquisition, testing intervals, visual field programs, and ocular comorbidities are less standardized.

AI-CO is commercially available clinical decision-support software that processes three-dimensional OCT image data and presents visual-function-related reference information. Its numerical outputs are unitless, and the manufacturer has not disclosed how the displayed numerical outputs are derived or calibrated. Their clinical interpretation therefore requires evaluation of how they relate to established functional measures.

We therefore externally evaluated the commercially implemented AI-CO system during its initial use at a community ophthalmology clinic. Our primary objectives were to quantify the rank association between the unitless output labeled “peripheral” in the AI-CO interface and Humphrey mean deviation (MD) and to assess discrimination of MD-defined severity thresholds. Additional analyses evaluated robustness in a fixed one-eye-per-patient cohort and across the 24-2 SITA Faster (24-2F) and 30-2 testing programs and estimated adjusted threshold-specific associations and severity probabilities.

Exploratory analyses evaluated nonlinearity, a severe-range floor effect, and cutoffs.

## Methods

### Study design and participants

This retrospective, single-center observational study was conducted in accordance with the Declaration of Helsinki and was approved by the Institutional Review Board of Jichi Medical University Hospital on August 21, 2026 (approval No. Rindai 26-068). Information about the study was publicly disclosed, and patients were given the opportunity to opt out.

Patients receiving care for glaucoma at Kora Eye Clinic were eligible. Glaucoma was diagnosed by ophthalmologists on the basis of optic disc appearance, OCT findings of the retinal nerve fiber layer and ganglion cell layer, visual field findings, and intraocular pressure; patients already receiving treatment were also included. During the initial implementation period, from April 2026 through the first week of May 2026, four orthoptists obtained OCT images as part of routine care. The candidate dataset comprised 824 eye records from 425 patients. After removal of two duplicate records and two eyes from a patient whose source examination did not meet the 365-day criterion on chart review, 820 eyes from 424 patients had successful AI-CO output and a corresponding Humphrey 24-2F or 30-2 examination within 365 days. One eye with a Humphrey MD of +19.83 dB and extensive false-positive responses was excluded because the visual field result was considered invalid. The final cohort comprised 819 eyes of 424 patients. To preserve routine-practice heterogeneity, no other uniform exclusion criterion based on visual field reliability indices or ocular comorbidities was applied.

### Visual field testing and OCT imaging

Humphrey visual field testing was performed with the Humphrey Field Analyzer (HFA) II-i or HFA 3 model 840 (Carl Zeiss Meditec). Eyes tested with the central 24-2 program using the SITA Faster strategy on the HFA3 840 were designated the 24-2F group. Eyes tested with the central 30-2 program using the SITA Fast strategy on either the HFA II-i or HFA3 840 were designated the 30-2 group. All examinations used a white Goldmann size III stimulus on a 31.5-asb background. OCT imaging was performed with a Topcon TRITON system using the 12×9-mm wide-scan mode. The interval was the absolute number of days between OCT imaging and perimetry; examinations before or after imaging were eligible within 365 days. For each AI-CO analysis, the eligible Humphrey examination closest in time to the OCT imaging date was selected, and no eye contributed to both program groups.

### AI-CO output

AI-CO (Software for Automated Visual Field and Ophthalmic Imaging [OCT], AI-CO; DeepEyeVision Co., Ltd.) is commercially available medical device software that imports three-dimensional OCT image data and displays a shaded planar grayscale image and a pattern-deviation-like image as visual-function-related reference information (Figure 1). All outputs were generated with AI-CO version 1.0.0. No data from Kora Eye Clinic were used to develop, train, or tune the AI-CO model. The Japanese interface displays two unitless numerical outputs labeled “peripheral” and “central”; their exact spatial definitions are not specified in the information available to the investigators. For consistency with the index name used in the clinic’s analyses, the output labeled “peripheral” in the Japanese interface is referred to as “AI-CO all” in this study; this name does not establish its anatomical coverage. The “central” output, termed AI-CO center in Figure 1, was outside the scope of this study.

**Figure 1.**
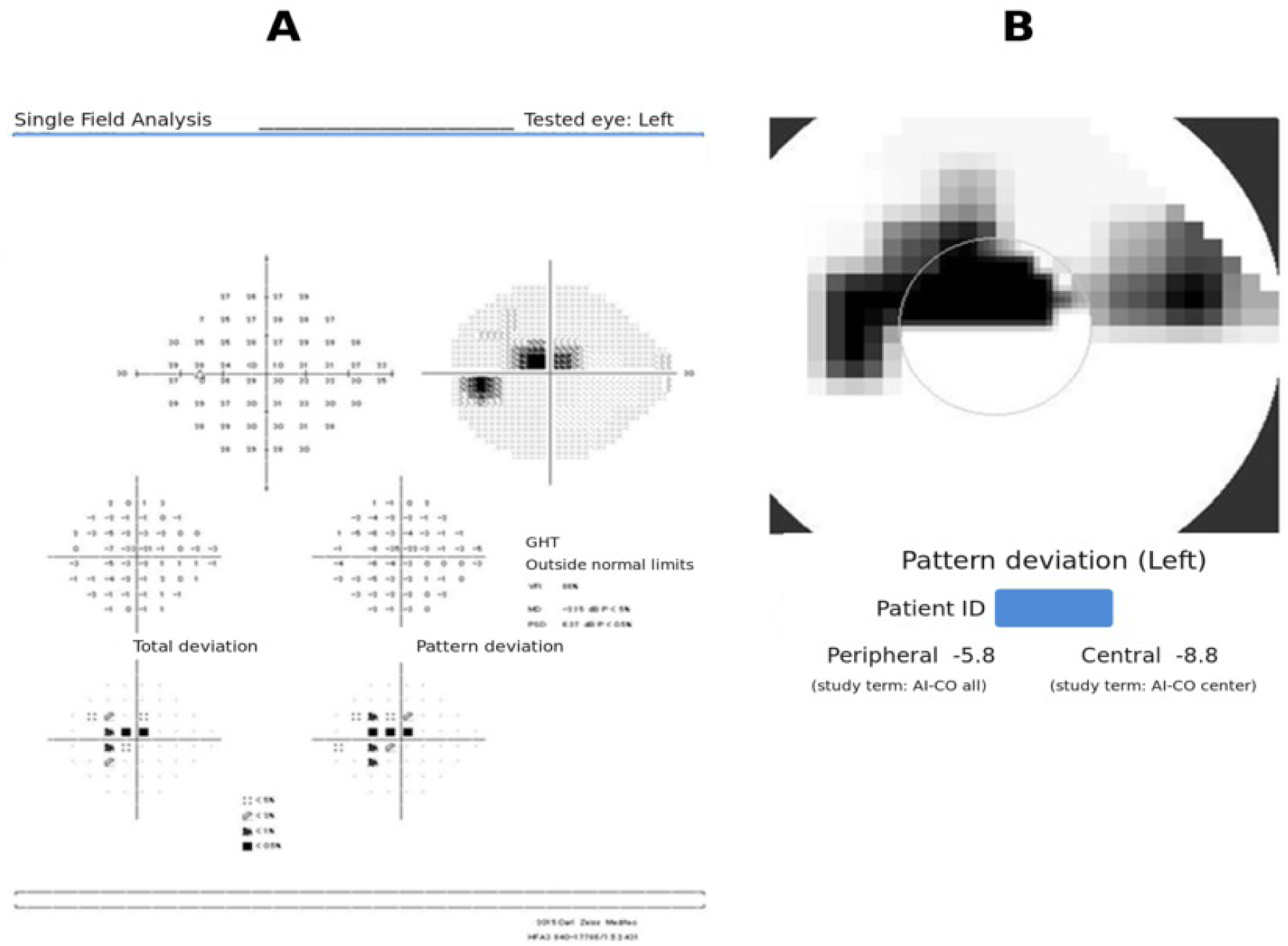
Example of AI-CO output and the corresponding Humphrey visual field examination. The Humphrey 24-2 visual field obtained with SITA Faster is shown in panel A, and the AI-CO pattern-deviation-like image from the same eye is shown in panel B. English translations of the Japanese interface labels are shown. Patient identifiers and the imaging date were masked.

### Severity definition and outcomes

Visual field severity was defined solely from Humphrey MD for the present analyses: normal range, MD ≥−2 dB; mild, −6 dB ≤MD <−2 dB; moderate, −12 dB ≤MD <−6 dB; and severe, MD <−12 dB. These categories were defined solely by MD for this study and do not constitute a comprehensive clinical staging system. The primary outcomes were the Spearman rank correlation between AI-CO all and Humphrey MD and the ability of AI-CO all to distinguish eyes across the MD thresholds of −2, −6, and −12 dB, quantified by the area under the receiver operating characteristic curve (AUC). Additional analyses evaluated severity-category distributions, covariate-adjusted threshold-specific associations and probabilities, and robustness across visual field programs and in a fixed one-eye-per-patient cohort.

Exploratory analyses evaluated cutoffs and internal validation, nonlinearity, and a severe-range floor effect.

### Statistical analysis

#### Rank association between AI-CO all and Humphrey MD

Spearman rank correlations were calculated overall and separately for the 24-2F and 30-2 groups. To account for correlation between fellow eyes, confidence intervals for the main eye-level analyses were estimated by resampling patients, with all eligible eyes of each selected patient kept together. This procedure was repeated 2,000 times. The difference between program-specific Spearman coefficients was evaluated with the same patient-level resampling method.

### AI-CO distributions across MD-defined severity categories

AI-CO all distributions across the four MD-defined severity categories were compared using the Kruskal–Wallis test. An ordered trend was evaluated by Spearman rank correlation between the ordinal severity category and AI-CO all.

### Discrimination of MD-defined severity

For each receiver operating characteristic (ROC) analysis, eyes with Humphrey MD below the specified threshold were defined as positive, and eyes with MD at or above the threshold were defined as negative. Thus, the positive outcomes were MD <−2 dB, MD <−6 dB, and MD <−12 dB. ROC analyses were oriented so that lower AI-CO values indicated greater impairment and a higher probability of the MD-positive outcome. Sensitivity was the proportion of MD-positive eyes correctly identified, and specificity was the proportion of MD-negative eyes correctly identified. AUC confidence intervals were obtained by repeating the patient-level resampling procedure 2,000 times. We also examined AUCs for adjacent severity categories and compared AUCs between the 24-2F and 30-2 groups.

### One-eye-per-patient sensitivity analysis

A sensitivity analysis repeated the rank correlation and three threshold AUCs in a fixed one-eye-per-patient cohort. For patients with two eligible eyes, one eye was selected using the saved fixed pseudorandom selection; patients with one eligible eye contributed that eye. The final selection comprised 424 eyes from 424 patients. The 95% confidence interval for Spearman correlation was calculated using the Fisher z approximation. The 95% confidence intervals for AUCs were computed from the 2.5th and 97.5th percentiles of 2,000 bootstrap resamples of the 424 selected eyes.

### Covariate-adjusted associations with MD-defined severity

To examine whether the association between AI-CO all and MD-defined severity persisted after accounting for routine-care differences, we fitted separate logistic regression models for each cumulative MD threshold. To assess whether a common odds ratio summarized the associations, patient-cluster robust Wald tests compared the coefficients from the three threshold-specific models: a global test compared all covariate slopes (10 degrees of freedom), and an index-specific test compared the AI-CO all slopes (2 degrees of freedom). AI-CO all was scaled per one standard deviation (SD) worsening (a 5.883-point decrease), and the models were adjusted for age, sex, visual field program, and interval between OCT imaging and perimetry. Confidence intervals and P values were calculated with patient-cluster robust standard errors to account for correlation between fellow eyes.

### Adjusted probabilities of MD-defined severity

Adjusted probabilities for the four severity categories were estimated using multinomial logistic regression with a restricted cubic spline for AI-CO all with knots at its 5th, 35th, 65th, and 95th percentiles and the same covariates. Predicted probabilities were averaged across all eyes while retaining each eye’s observed age, sex, visual field program, and examination interval.

### Exploratory AI-CO cutoffs and internal validation

Exploratory cutoffs were selected by maximizing the Youden index. To examine their performance in data not used to select them, patients were divided into 10 groups. All eligible eyes from the same patient were kept in the same group to prevent information from the same patient from appearing in both the training and test data. The cutoff was determined using nine groups and evaluated in the remaining group. This process was repeated until each group had served as the test group, and the entire 10-fold procedure was repeated 50 times.

### Continuous relationship and nonlinearity

To explore the continuous relationship, we fitted unadjusted and covariate-adjusted linear and restricted cubic spline models of AI-CO all against Humphrey MD. Spline knots were placed at the 5th, 35th, 65th, and 95th percentiles of MD. Patient-cluster robust standard errors were used; the nonlinear spline terms were tested jointly with a Wald test.

### Assessment of a severe-range floor effect

Local slopes were obtained as derivatives of the fitted adjusted spline with respect to MD, with patient-cluster robust 95% confidence intervals. The slope is constant from MD −20 to −25 dB because this interval lies below the first spline knot (MD −17.46 dB). The number of eyes at the observed minimum AI-CO all value was also examined for evidence of a severe-range floor effect.

### Statistical inference and software

All tests were two-sided, with P<0.05 considered statistically significant. Rank correlations, AUCs, one-eye Spearman confidence interval by Fisher z approximation and one-eye AUC bootstrap confidence intervals, group comparison tests, threshold-specific models, spline models, and multinomial probabilities were calculated or verified in Python 3.12 (NumPy 2.3.5, SciPy 1.17.0, and scikit-learn 1.8.0).

## Results

### Participants

The analysis included 819 eyes of 424 patients: 343 eyes in the 24-2F group and 476 eyes in the 30-2 group. Mean age was higher in the 30-2 group than in the 24-2F group, and the imaging-to-perimetry interval was longer in the 24-2F group (both P<0.001). Sex distribution and Humphrey MD did not differ significantly between the groups (Table 1).

**Table 1.** Characteristics of the visual field program groups.

| Characteristic | 24-2F (n=343) | 30-2 (n=476) | P value |
| --- | --- | --- | --- |
| Age, years | 66.98 ± 13.69 | 71.00 ± 10.80 | <0.001 |
| Sex | Male 148 (43.1%)<br>Female 195 (56.9%) | Male 206 (43.3%)<br>Female 270 (56.7%) | 0.971 |
| Humphrey MD, dB | -5.07 ± 5.31 | -5.64 ± 6.09 | 0.154 |
| Examination interval, days | 83 (0–181) | 1 (0–119) | <0.001 |

Values are mean ± standard deviation, n (%), or median (interquartile range). Age and MD were compared by Welch t test, sex by Pearson chi-square test, and examination interval by Mann–Whitney U test. MD = mean deviation.

### Rank association between AI-CO and Humphrey MD

AI-CO all was correlated with Humphrey MD in the full cohort (Spearman ρ=0.785; patient-cluster 95% CI, 0.750–0.818; P<0.001). Correlations were similar in the 24-2F group (ρ=0.784; 95% CI, 0.730– 0.830) and the 30-2 group (ρ=0.786; 95% CI, 0.739–0.827). Their difference was −0.002 (95% CI, −0.075 to 0.062; P=0.940).

### AI-CO distributions across MD-defined severity categories

When eyes were grouped according to Humphrey MD, AI-CO distributions progressively shifted toward lower values with increasing severity (Table 2 and Figure 2). The distributions differed across the four groups, and the ordered decrease was statistically significant (both P<0.001).

**Figure 2.**
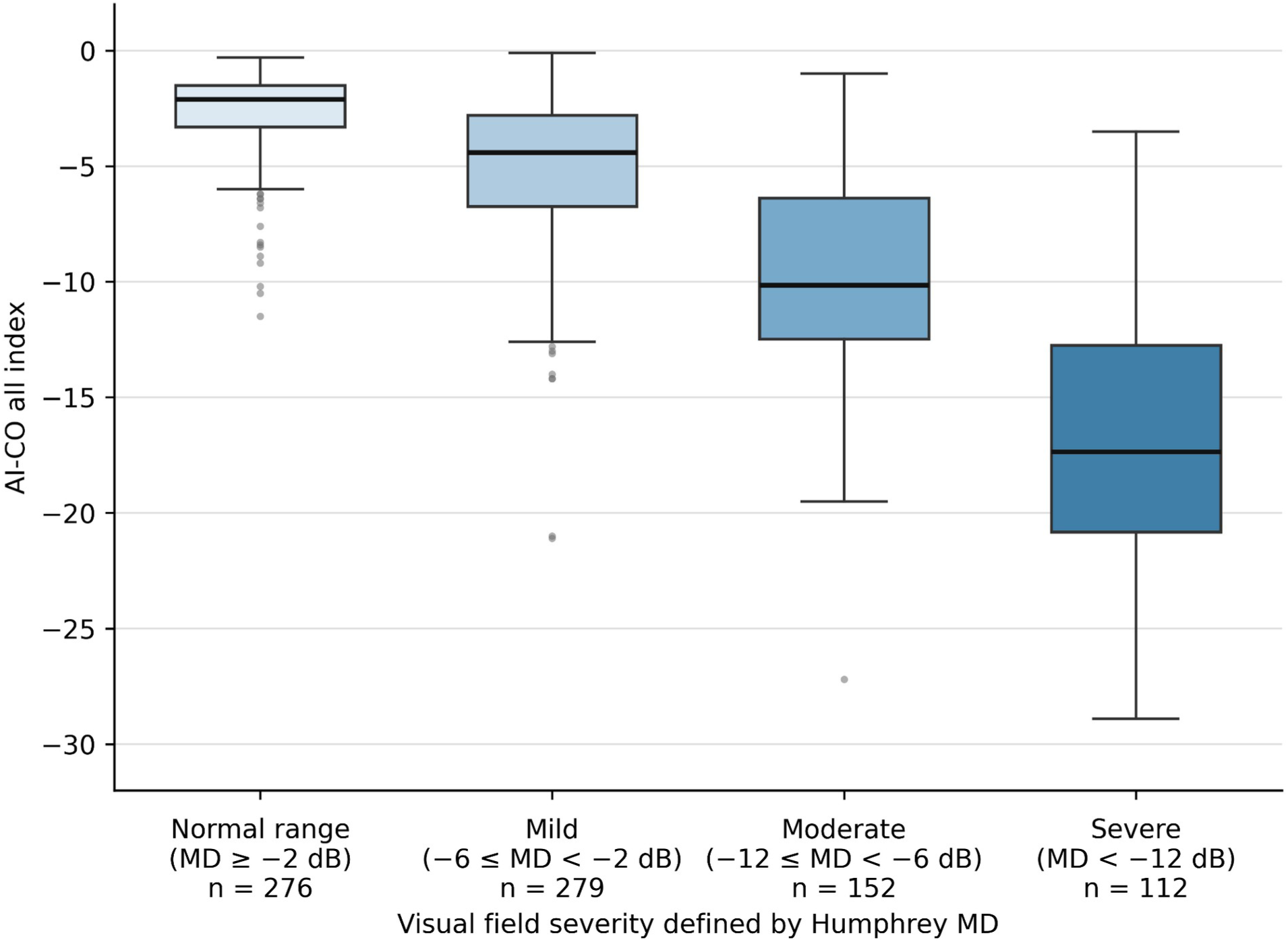
Distribution of AI-CO values across MD-defined severity categories. Boxes represent the interquartile range, horizontal lines indicate medians, whiskers extend to 1.5 times the interquartile range, and circles indicate values beyond the whiskers. Severity was defined solely by Humphrey MD. Median AI-CO all values decreased monotonically with increasing MD-defined severity.

**Table 2.** AI-CO distribution by MD-defined severity.

| Severity category | MD definition | n | Median AI-CO | Interquartile range |
| --- | --- | --- | --- | --- |
| Normal range | ≥-2 dB | 276 | -2.10 | -3.30 to -1.50 |
| Mild | -6 to <-2 dB | 279 | -4.40 | -6.75 to -2.80 |
| Moderate | -12 to <-6 dB | 152 | -10.15 | -12.48 to -6.38 |
| Severe | <-12 dB | 112 | -17.35 | -20.83 to -12.75 |

Kruskal–Wallis P<0.001; monotonic trend P<0.001. Severity strata were defined solely by Humphrey MD and do not constitute a comprehensive clinical staging system.

### Discrimination of MD-defined severity

AI-CO all differentiated eyes across all three cumulative severity thresholds (Table 3 and Supplementary Figure S1). The AUC was 0.870 (95% CI, 0.842–0.895) for MD <−2 dB, 0.919 (95% CI, 0.894–0.939) for MD <−6 dB, and 0.943 (95% CI, 0.920–0.963) for MD <−12 dB. Patient-cluster bootstrap comparisons showed no significant difference in program-specific AUCs between 24-2F and 30-2 at any threshold (Supplementary Table S1).

**Table 3.** Discrimination of cumulative MD severity thresholds.

| Positive outcome | Positive eyes | AUC | Patient-cluster 95% CI |
| --- | --- | --- | --- |
| MD <-2 dB | 543 | 0.870 | 0.842–0.895 |
| MD <-6 dB | 264 | 0.919 | 0.894–0.939 |
| MD <-12 dB | 112 | 0.943 | 0.920–0.963 |

Discrimination remained present when only adjacent categories were compared: AUCs were 0.779 for normal range versus mild, 0.811 for mild versus moderate, and 0.824 for moderate versus severe (Supplementary Table S2). Thus, the cumulative-threshold performance was not explained solely by separation of normal-range from severely affected eyes.

### One-eye-per-patient sensitivity analysis

In the fixed one-eye-per-patient cohort of 424 eyes, the Spearman correlation was 0.781 (95% CI, 0.740 –0.815). AUCs were 0.878, 0.920, and 0.940 for the three cumulative thresholds, closely matching the main analysis (Supplementary Table S3).

### Covariate-adjusted associations with MD-defined severity

The proportional-odds assumption was not fully supported (global P=0.042), and the AI-CO all association differed across the three cumulative MD thresholds (P=0.0066). In separate threshold-specific models adjusted for age, sex, visual field program, and examination interval, a 1-SD lower AI-CO value was associated with higher odds of mild-or-worse versus normal-range MD (adjusted OR, 34.61; 95% CI, 15.85–75.56), moderate-or-worse versus mild-or-better MD (adjusted OR, 13.95; 95% CI, 9.24–21.04), and severe versus moderate-or-better MD (adjusted OR, 8.79; 95% CI, 5.84–13.23); all P<0.001 (Table 4).

**Table 4.** Adjusted association between AI-CO worsening and MD-defined severity.

| Cumulative comparison | Adjusted OR per 1-SD lower AI-CO | 95% CI | P value |
| --- | --- | --- | --- |
| Mild or worse vs normal range | 34.61 | 15.85–75.56 | <0.001 |
| Moderate or worse vs mild or better | 13.95 | 9.24–21.04 | <0.001 |
| Severe vs moderate or better | 8.79 | 5.84–13.23 | <0.001 |

Separate threshold-specific logistic models were adjusted for age, sex, visual field program, and examination interval, with patient- cluster robust standard errors. One SD corresponded to a 5.883-point decrease in AI-CO. OR = odds ratio.

### Adjusted probabilities of MD-defined severity

Standardized predicted probabilities varied by AI-CO all value (Figure 3 and Supplementary Table S4). At an AI-CO value of −2, the predicted probability of normal-range MD was 70.6%; at −5, mild severity had the highest probability (60.6%); at −10, moderate severity had the highest probability (49.4%); and at −20, the severe category had a predicted probability of 83.2%. At −15, moderate and severe probabilities overlapped (44.4% and 46.1%, respectively), illustrating that the index does not assign an individual eye to a deterministic category.

**Figure 3.**
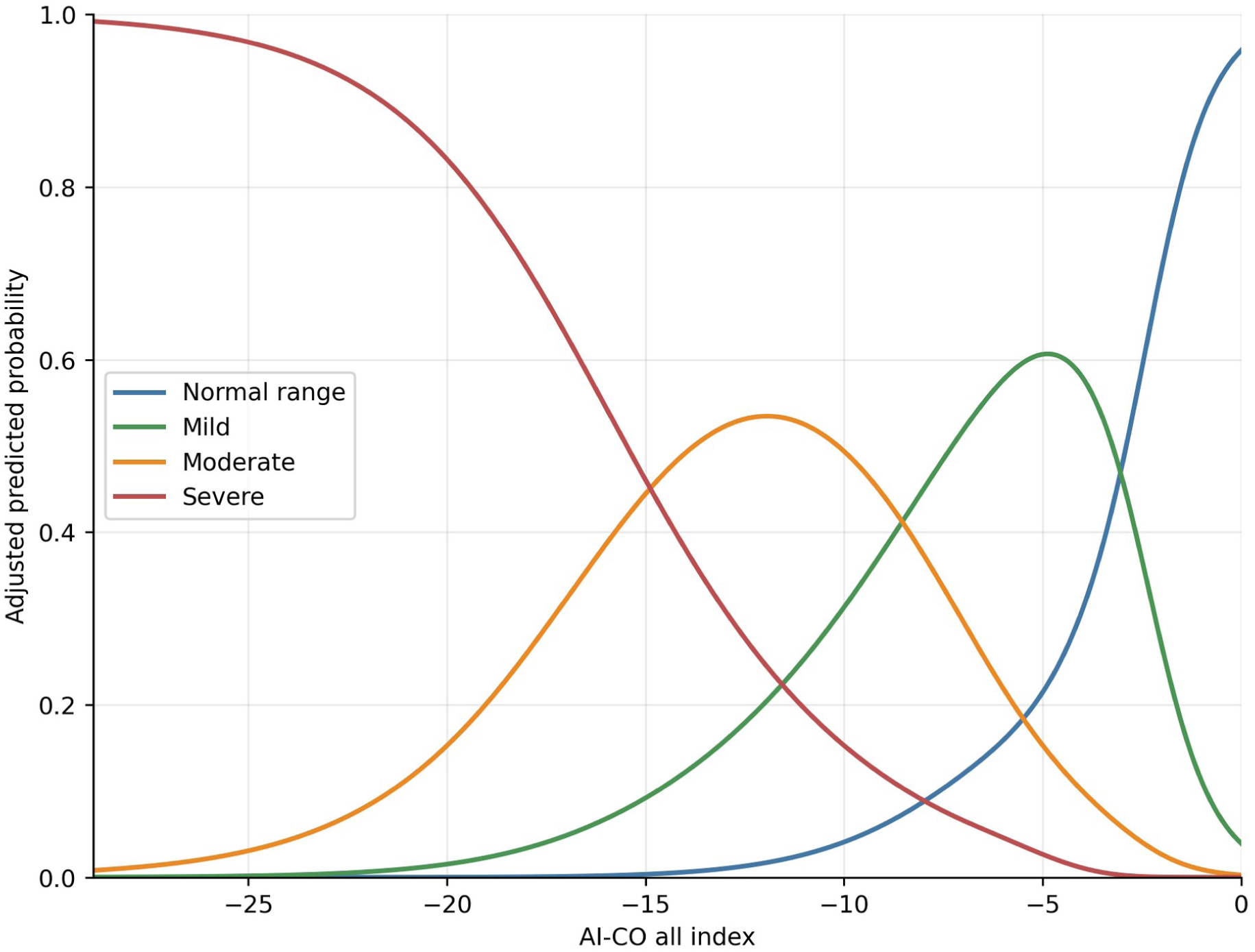
Adjusted predicted probabilities of MD-defined severity by AI-CO all value. Probabilities were obtained from multinomial logistic regression with a restricted cubic spline for AI-CO all and adjustment for age, sex, visual field program, and examination interval. Predictions retained each eye’s observed covariate values and were then averaged across all eyes.

### Exploratory AI-CO cutoffs and internal validation

In repeated patient-level cross-validation, balanced accuracy was 0.769–0.866 (Supplementary Table S5). The estimated cutoffs and their sensitivity and specificity remain exploratory and are not clinical decision thresholds.

### Continuous relationship and nonlinearity

The adjusted continuous spline model detected modest nonlinearity (P=0.00081), but the increase in explained variance over the linear model was small (R^2^, 0.735 to 0.738; Supplementary Table S6 and Figure 4). The fitted curve showed a shallower local slope near normal MD and retained a positive slope across the observed severe range, without an abrupt break in the association. Above the upper spline knot (MD 0.471 dB), the fitted slope remained positive (0.379; 95% CI, 0.148–0.609) across 41 eyes from 36 patients.

**Figure 4.**
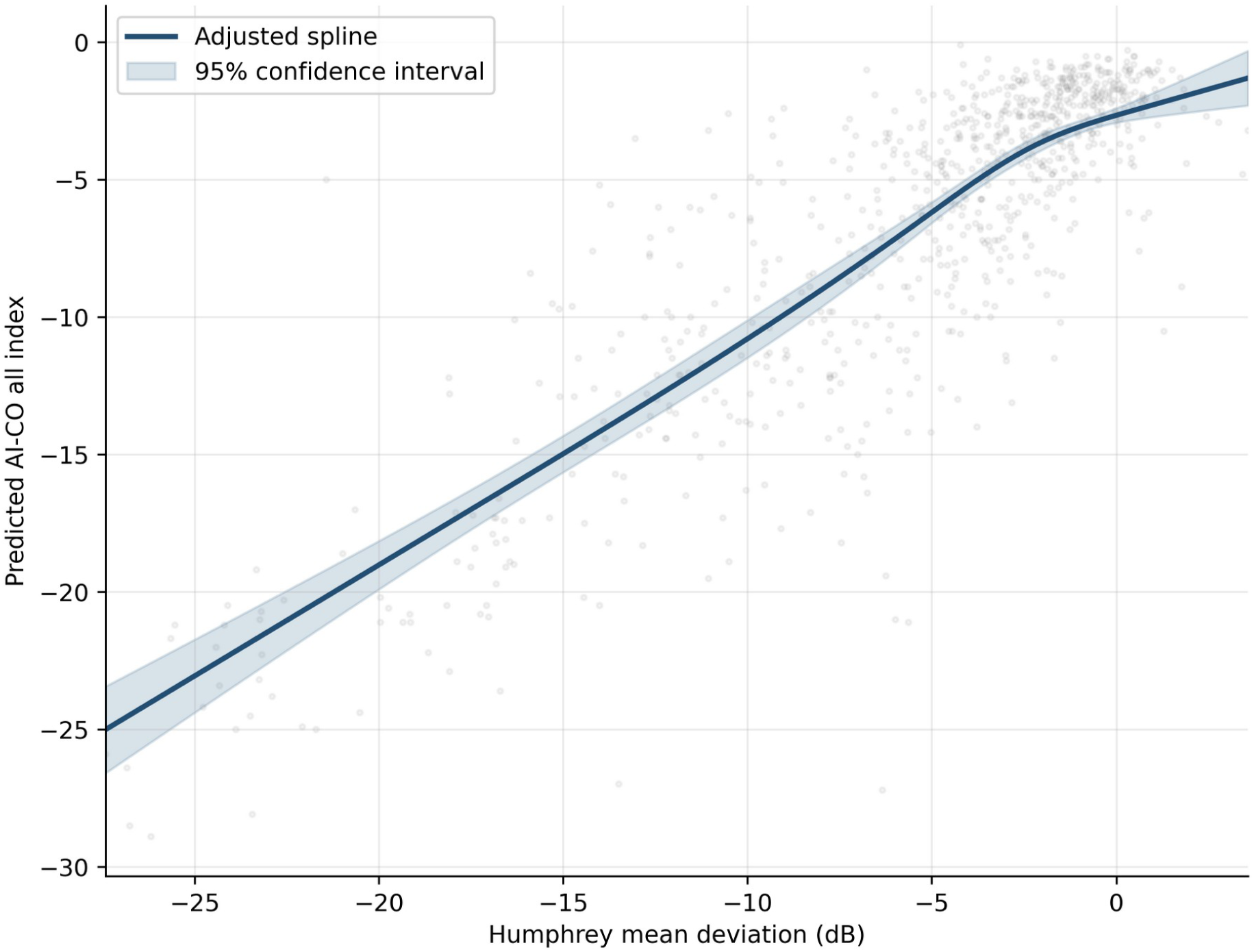
Adjusted restricted cubic spline relationship between Humphrey MD and AI-CO all. Gray points indicate individual observed eyes; the solid curve shows the adjusted predicted AI-CO value at the cohort mean covariate values and the shaded region shows the 95% confidence interval. The nonlinear component was statistically significant, but the increase in R^2^ compared with the linear model was 0.003.

### Assessment of a severe-range floor effect

Local slopes remained positive at the severe boundary and across the observed severe range: 0.841 (95% CI, 0.764–0.917) at MD −12 dB and 0.808 (95% CI, 0.692–0.923) at MD −20 to −25 dB. The minimum AI-CO value of −28.9 occurred in only one eye. These findings did not support a clear severe-range floor effect, although a device-defined theoretical lower bound was unavailable (Supplementary Table S6).

## Discussion

In this independent real-world evaluation of a commercially implemented system, AI-CO all showed a Spearman rank correlation of 0.785 with Humphrey MD and differentiated eyes across specified MD thresholds. Similar findings were observed in the 24-2F and 30-2 groups, in the fixed one-eye-per-patient cohort, and after adjustment for age, sex, visual field program, and examination interval. These findings support an association between AI-CO values and MD-defined severity under the routine-care conditions evaluated in this study.

Humphrey MD is expressed in decibels, whereas AI-CO all is unitless and the derivation and calibration of its displayed numerical values have not been disclosed. As an OCT-derived index, AI-CO should be interpreted as providing severity-related information that complements measured perimetry. Lower AI-CO values were associated with worse MD and differentiated MD-defined severity categories (Tables 2 and 3; Figures 2 and 4).

Overlap in AI-CO values across MD-defined severity categories should be interpreted in the context of fundamentally different measurement approaches. Standard automated perimetry measures visual function through patient responses, whereas AI-CO derives severity-related information from structural OCT data. Humphrey MD served as the clinical reference for defining severity in this study, but not as an error-free biological ground truth. The observed overlap may therefore reflect perimetric variability, structure–function discordance, differences in the information captured by each method, or model-related estimation error (Figure 2).

Previous studies have shown that deep-learning models can estimate visual field sensitivity or global indices from OCT-derived data^1–6^. The present study extends that work by independently evaluating a deployed commercial system using routine-care images from a community clinic, two visual field workflows, and clinically heterogeneous eyes. It also accounts for correlation between fellow eyes and evaluates severity discrimination and adjusted severity probabilities rather than relying on correlation alone. The adjacent-category analyses further showed that discrimination was not produced only by contrasting the least and most severely affected eyes (Supplementary Table S2).

The adjusted OR per one SD lower AI-CO all value was largest at the mild-or-worse threshold and smaller at the more severe thresholds (Table 4), indicating that the relationship between AI-CO and MD-defined severity was not adequately summarized by a single common odds ratio.

The continuous association across the observed MD range was approximately linear, with a small but statistically detectable nonlinear component and only a small improvement in explained variance from spline modeling (Figure 4). Although the nonlinear component was statistically detectable, the spline depicts a continuous relationship rather than a marked change in direction between severity levels. Its shallower local slope near normal MD suggests compression at the less impaired end, but this should not be labeled a ceiling effect because the theoretical bounds of the proprietary index are unknown.

Conversely, preserved local slopes and absence of accumulation at the observed minimum argue against a clear floor effect in the severe range. These exploratory findings may be relevant when designing longitudinal studies, but they require confirmation with repeated measurements.

Predicted probabilities describe how selected AI-CO values were distributed across severity categories, but the overlap at intermediate values is important: AI-CO does not provide a deterministic diagnosis or substitute for measured perimetry in an individual eye.

Exploratory AI-CO all cutoffs were evaluated with patient-level cross-validation, but the cutoffs were derived and evaluated within the same single-center population. Independent cohorts are needed before any cutoff is considered for clinical application.

Several limitations should be considered. The study was retrospective and conducted at a single center. AI-CO imaging and Humphrey perimetry were not necessarily performed on the same day, and only one OCT device and scan range were evaluated. The 30-2 group included two perimeter models. Eligibility required successful AI-CO output, so imaging success and applicability to all routine-care patients could not be estimated. Severity categories were defined only by MD and were not a complete staging system. Conventional OCT thickness indices were not available for direct comparison, preventing assessment of incremental discrimination beyond standard structural measures. The proprietary scale and spatial definitions of the AI-CO outputs were unavailable. Finally, the cutoffs and severity probabilities were derived internally and require validation in an independent population.

The present findings do not support replacing standard automated perimetry. AI-CO may provide complementary structural-functional information when contemporaneous perimetry is unavailable and may help prioritize formal functional testing; these potential uses were not evaluated in this study, and clinical utility, effects on decision-making, longitudinal responsiveness, and patient outcomes require prospective evaluation.

## Conclusions

In this real-world external evaluation, AI-CO all showed a rank correlation with Humphrey MD and differentiated eyes across the specified MD thresholds. Similar findings were obtained after accounting for inclusion of both eyes, in the one-eye-per-patient sensitivity analysis, across visual field programs, and after adjustment for age, sex, visual field program, and examination interval. The continuous association was approximately linear, with small but statistically detectable nonlinearity and no clear evidence of a severe-range floor effect. AI-CO should be interpreted as an OCT-derived severity-related index that complements, rather than replaces, measured perimetry. Exploratory AI-CO all cutoffs require independent validation before clinical application.

## Conflict of interest statement

AI-CO was initially made available to Kora Eye Clinic for a clinical demonstration and was subsequently purchased by the clinic through an ordinary commercial transaction. The authors received no research funding, data analysis, or other analytical support for this study from DeepEyeVision Co., Ltd. Y.K. is scheduled to receive an honorarium from DeepEyeVision Co., Ltd. for a scientific lecture that includes presentation of some findings from this study. DeepEyeVision Co., Ltd. had no role in the study design, conduct, data collection, analysis, interpretation, manuscript preparation, or decision to submit the manuscript for publication. The other authors declare no competing interests relevant to this work.

## Funding

This study received no external funding.

## Acknowledgments

The authors thank the orthoptists at Kora Eye Clinic for their assistance with data collection. During the preparation of this manuscript in September 2026, the authors used GPT-5.6 Sol and GPT-6 through ChatGPT/Codex (OpenAI, https://chatgpt.com) to assist with statistical coding, verification of analytical procedures and numerical consistency, generation of data visualizations, organization and drafting of manuscript content, and English-language editing. Claude Sonnet 5 and Claude Opus 5.5 through Claude (Anthropic, https://claude.ai) were used to review the manuscript for internal consistency and clarity. All analytical methods, interpretations, and final editorial decisions were made and approved by the authors. The authors reviewed the analyses, figures, references, interpretations, and manuscript text and take responsibility for the submitted work.

## Author contributions

Y.K.: Conceptualization, Methodology, Formal analysis, Investigation, Data curation, Visualization, Writing – original draft, and Project administration. H.K.: Investigation, Data curation, Validation, and Writing – review and editing. T.K.: Investigation, Data curation, Validation, and Writing – review and editing. All authors reviewed and approved the final manuscript.

## Data availability statement

The de-identified data that support the findings of this study are available from the corresponding author upon reasonable request, subject to institutional ethical and privacy requirements.

## Funding

None

## Conflict of interest

Y.K. is scheduled to receive an honorarium from DeepEyeVision Co., Ltd. for a scientific lecture that includes presentation of some findings from this study. The company had no role in the study or manuscript preparation. H.K. and T.K. declare no competing interests relevant to this work.

## Supplementary tables

**Supplementary Table S1.**
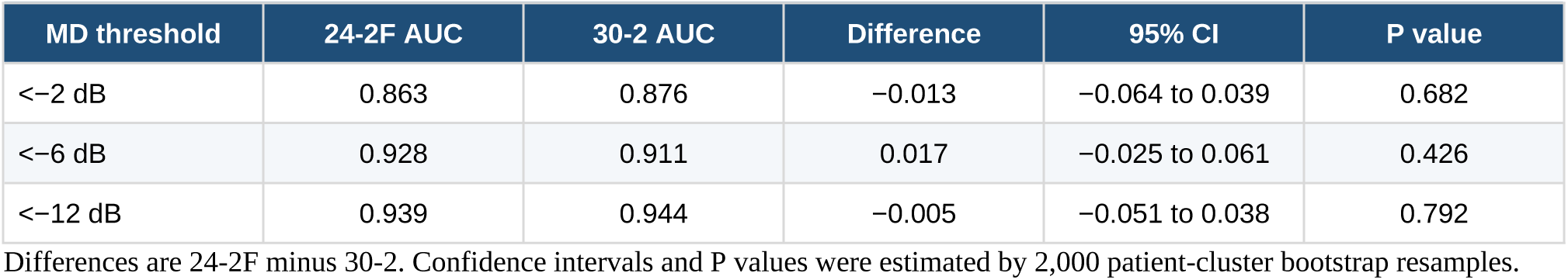
Program-specific AUCs and between-program differences.

**Supplementary Table S2.** Discrimination between adjacent severity categories.

| Comparison | n | AUC | Patient-cluster 95% CI |
| --- | --- | --- | --- |
| Normal range vs mild | 555 | 0.779 | 0.739–0.819 |
| Mild vs moderate | 431 | 0.811 | 0.765–0.854 |
| Moderate vs severe | 264 | 0.824 | 0.769–0.880 |

**Supplementary Table S3.** Fixed one-eye-per-patient sensitivity analysis.

| Analysis | n | Spearman $\rho$ or AUC | 95% CI |
| --- | --- | --- | --- |
| Spearman correlation | 424 | 0.781 | 0.740–0.815 |
| AUC for MD <-2 dB | 424 | 0.878 | 0.846–0.908 |
| AUC for MD <-6 dB | 424 | 0.920 | 0.892–0.945 |
| AUC for MD <-12 dB | 424 | 0.940 | 0.908–0.966 |

The fixed one-eye-per-patient selection used previously saved random numbers, as described in Methods. The Spearman correlation confidence interval used the Fisher z approximation; AUC confidence intervals used 2,000 bootstrap resamples of the selected eyes.

**Supplementary Table S4.** Adjusted predicted severity probabilities at selected AI-CO values.

| AI-CO | Normal range | Mild | Moderate | Severe |
| --- | --- | --- | --- | --- |
| -2 | 70.6% | 26.7% | 2.6% | 0.03% |
| -5 | 21.4% | 60.6% | 15.3% | 2.7% |
| -10 | 4.1% | 31.3% | 49.4% | 15.3% |
| -15 | 0.3% | 9.2% | 44.4% | 46.1% |
| -20 | 0.01% | 1.5% | 15.2% | 83.2% |

Probabilities are standardized average marginal predictions from a multinomial logistic model with restricted cubic spline terms for AI-CO and adjustment for age, sex, visual field program, and examination interval. Rounding may prevent totals from equaling exactly 100%. Spline knots were at the 5th, 35th, 65th, and 95th percentiles of AI-CO all (-20.2, -7.1, -3.1, and -1.1).

**Supplementary Table S5.**
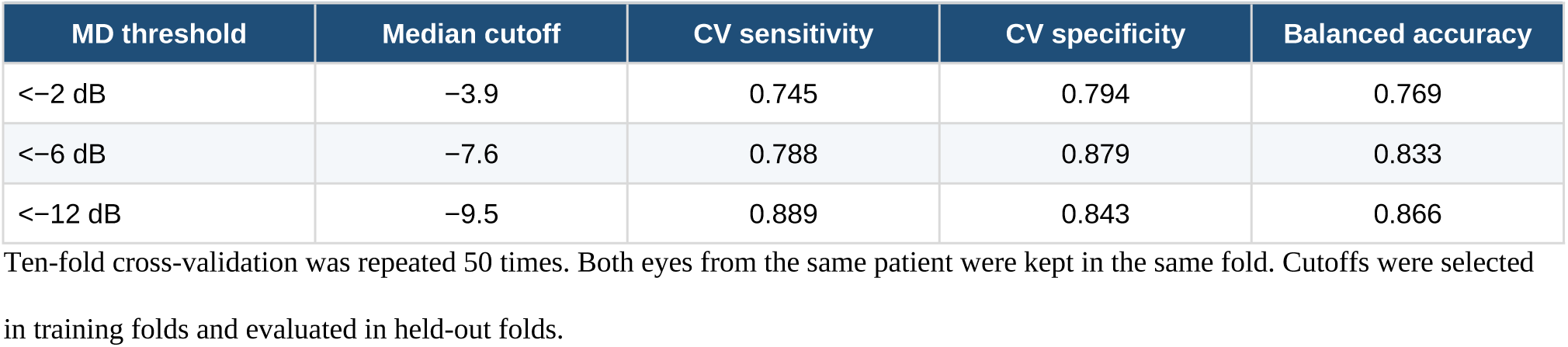
Repeated patient-level cross-validation of exploratory cutoffs.

**Supplementary Table S6.** Continuous nonlinearity and local slope assessment.

| Analysis or MD value | Linear R<sup>2</sup> | Spline R<sup>2</sup> | Nonlinearity P | Local slope | 95% CI |
| --- | --- | --- | --- | --- | --- |
| Unadjusted model | 0.719 | 0.722 | 0.0021 | — | — |
| Adjusted model | 0.735 | 0.738 | 0.00081 | — | — |
| MD >0.471 dB | — | — | — | 0.379 | 0.148–0.609 |
| MD 0 dB | — | — | — | 0.388 | 0.162–0.613 |
| MD -2 dB | — | — | — | 0.629 | 0.517–0.740 |
| MD -6 dB | — | — | — | 0.954 | 0.822–1.086 |
| MD -12 dB | — | — | — | 0.841 | 0.764–0.917 |
| MD -20 to -25 dB | — | — | — | 0.808 | 0.692–0.923 |

Adjusted models included age, sex, visual field program, and examination interval. Inference used patient-cluster robust standard errors. Local slope is $d(AI-CO)/d(MD)$ . Spline knots were at the 5th, 35th, 65th, and 95th percentiles of MD (-17.457, -5.391, -2.170, and 0.471 dB). Above the upper knot, the local slope is constant; 41 eyes from 36 patients had MD >0.471 dB.

## Supplementary figure legends

**Supplementary Figure S1.**
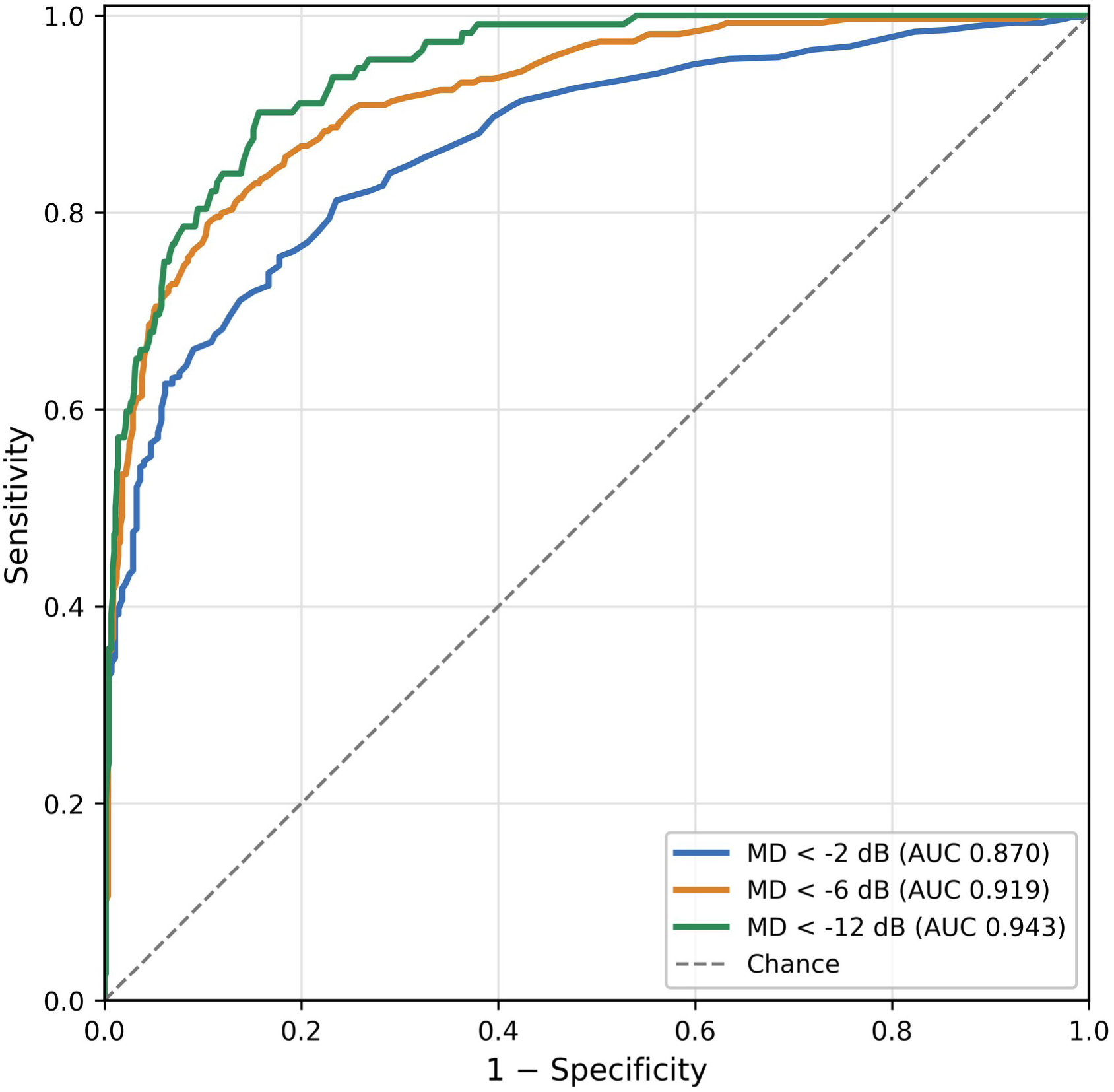
Receiver operating characteristic curves for cumulative MD severity thresholds. Curves show discrimination of MD <−2, <−6, and <−12 dB using AI-CO all, oriented so that lower AI-CO values indicated greater impairment. AUCs were 0.870, 0.919, and 0.943, respectively (patient-cluster bootstrap 95% CIs in Table 3).

## Notes

### Competing Interest Statement

After an initial demonstration period, AI-CO was purchased by our clinic through an ordinary commercial transaction. The authors received no research funding or analytical support related to this study from DeepEyeVision Inc. Y.K. is scheduled to receive an honorarium from DeepEyeVision Inc. for a scientific lecture. The other authors declare no competing interests relevant to this work.

### Author Declarations

The Institutional Review Board of Jichi Medical University Hospital gave ethical approval for this work (approval No. Rindai 26-068).

### Summary of Updates

This version substantially revises the analysis and interpretation of AI-CO output. Because the AI-CO all index is unitless, the manuscript no longer interprets its numerical values as interchangeable with Humphrey mean deviation in dB. The revised primary analyses assess rank association with mean deviation and discrimination of mean-deviation-defined visual field severity. Additional analyses examine adjacent severity categories, associations adjusted for clinical covariates, and robustness using one eye per patient and separate visual field programs. Exploratory analyses examine potential nonlinearity, severe-range behavior, and internally evaluated cutoffs. The title, abstract, methods, results, discussion, figures, tables, and supplementary material have been updated accordingly.

